# Immune (dys)function in obsessive-compulsive disorder

**DOI:** 10.1101/2025.04.23.25326223

**Authors:** Ana Maia, Nelson Descalço, Sara Fernandes, Catarina Fonseca, Tiago Quendera, Pedro Castro-Rodrigues, Fernanda Marques, Ana Daniela Costa, Pedro Morgado, Bernardo Barahona-Corrêa, José Oliveira, Albino J. Oliveira-Maia

## Abstract

**Background:** It has been suggested that immune-mediated neuronal damage plays a decisive role in the etiology of obsessive-compulsive disorder (OCD). Here we assessed an extensive panel of immune markers in the peripheral blood of two samples of patients with OCD and matched controls, recruited in distinct geographical areas.

**Methods:** Participants were assessed cross-sectionally for sociodemographic and clinical characteristics. High-sensitivity C-reactive protein, twelve cytokines, anti-nuclear, anti-thyroid peroxidase, anti-thyroglobulin, and anti-basal ganglia antibodies were assessed using ELISA. Cytokine gene expression was assessed in one of the populations using RT-qPCR.

**Results:** While patients had a significantly higher prevalence of self-reported general medical disorders, we consistently found similar systemic immune indicators between patients (n=139) and controls (n=131), across markers and populations.

**Discussion:** Our findings support that, contrary to what has been consistently shown in mood and psychotic disorders, in OCD there are no changes in peripheral immune function, when assess cross-sectionally in symptomatic adults.

## 1. Introduction

Mental disorders remain among the top leading causes of disease burden worldwide, with no evidence of global reduction^1^. This is the case of obsessive-compulsive disorder (OCD), a psychiatric disorder most commonly characterized by an early-onset and chronic course^2^, highly comorbid with both physical and other psychiatric conditions^3^, and associated with significant reductions of functionality and quality of life^4^. Notwithstanding the development of numerous evidence-based treatments for OCD, there are still high rates of treatment non-response and non-adherence, in part due to stigma and low accessibility. However, more effective strategies for prevention and treatment are also needed, and may be facilitated from enhanced understanding of the causes of OCD^5,6^. In recent years, there has been a growing interest on a potential role for the immune system in the etiology and pathophysiology of several psychiatric disorders, mainly mood disorders and psychosis ^7^. The most cited models propose that environmental factors trigger chronic immune dysfunction in genetically susceptible individuals^8^, with consistent empirical support for the association of these disorders with markers of abnormal immune homeostasis in the central nervous system (CNS) and the peripheral blood^9,10^.

The hypothetical role of immune dysregulation and chronic inflammation has also been considered for OCD and was initially fueled by the well-known association between obsessive-compulsive (OC) symptoms and recent infection with group A streptococcus in children, as part of a syndrome known as Pediatric Autoimmune Neuropsychiatric Disorder Associated with Streptococcal infections (PANDAS)^11^. This and later observations that OC symptoms often develop acutely in children following infection with other microorganisms have been seen as evidence that infections might act as an environmental risk factor for primary idiopathic OCD, through mechanisms possibly involving cross-reactivity of anti-microorganism antibodies with brain epitopes^12^. In fact, patients with OCD have an increased prevalence of immune-mediated physical illnesses^13–15^ and are often positive for anti-basal-ganglia antibodies (ABGA)^16^ in the peripheral blood. Moreover, one study using positron emission tomography showed increased translocator protein density, an indirect marker of microglia activation, particularly in the cortico-striato-thalamo-cortical circuit in unmedicated patients with OCD^17^. On the other hand, a single study assessing cytokines in the cerebrospinal fluid found similar interleukin (IL)-6 concentrations in patients and healthy volunteers^18^, while for systemic biomarkers evidence has been equivocal and contradictory. Indeed, available evidence suggests that patients with OCD do not have a higher prevalence of systemic general autoantibodies^19,20^, and two meta-analyses assessing cytokine concentration in peripheral blood showed conflicting results^21,22^.

In summary, although there is evidence suggesting that adult patients with OCD have a compromised systemic immune function characterized by altered markers of peripheral inflammation and autoimmunity^23^, studies supporting this claim obtained inconsistent results, mostly based on small samples and heterogeneous methodologies^19–22^. In this work, we conducted a cross-sectional observational study to compare a large sample of adult symptomatic patients with OCD and age- and sex-matched healthy volunteers, for peripheral markers of inflammation and auto-antibodies. To overcome limitations associated with recruitment and sample size, we reproduced our methods in a second independent sample, to assess if results were replicated.

## 2. Results

### a. Sample recruitment and description

One hundred and sixty-nine patients with a diagnosis of OCD were contacted between October 2019 and May 2023 in three clinical centers in Lisbon. Concomitantly, 201 controls without history of psychiatric disorders were identified in the general population in the same geographical area, amounting to a total of 369 subjects. After assessing eligibility criteria, 92 patients and 95 age and sex-matched controls were included and assessed at the Champalimaud Foundation (CF) in Lisbon (Portugal) (**Figure 1**).

**Figure 1.**
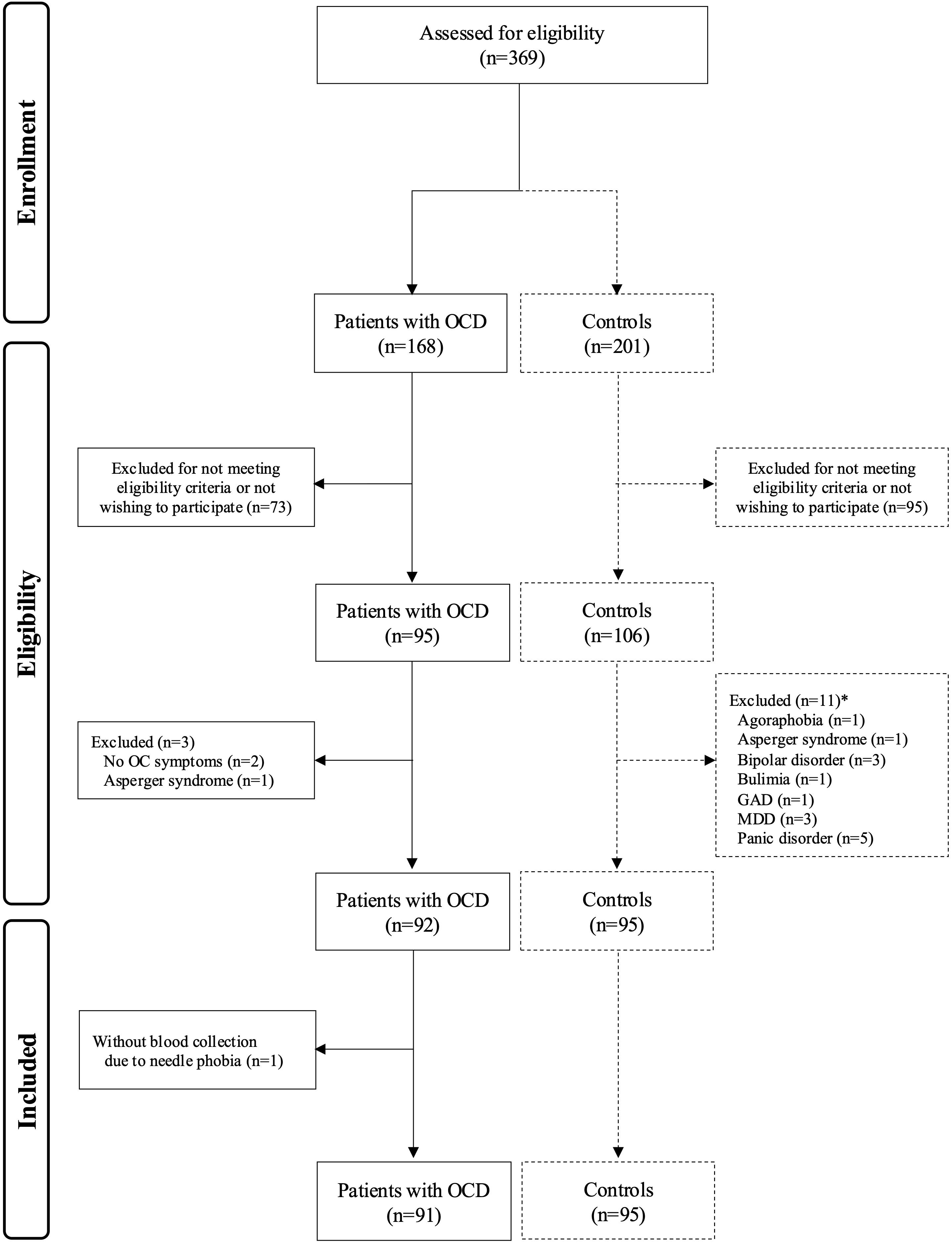
Recruitment flowchartI. **Legend:** GAD - generalized anxiety disorder; MDD-major depressive disorder; OC - obsessive-compulsive; OCD - obsessive-compulsive

In accordance with matching, patients and controls did not differ significantly regarding age and sex, and differences in body mass index (BMI) and percentage of smokers were also not found. On average, patients had slightly but significantly less years of education when compared with controls. Regarding clinical characteristics, patients had a mean age of 16.6±9.8 years at OCD onset, with 73.9% presenting an early-onset subtype (i.e. ≤19 years^24^), and with moderate illness severity on average (total Yale-Brown Obsessive-Compulsive Scale-I (YBOCS)-I score 20.0±6.3)^25^ when recruited. Patients had significantly more severe symptoms of depression and anxiety, when compared to controls, and almost one-third had a current depressive episode. Regarding pharmacological treatment, 76 (84.4%) of patients were medicated, and only one healthy control was taking psychotropic medication for insomnia. Fourteen (15.6%) patients were medication-naïve **(Table 1**).

**Table 1.** Sociodemographic and clinical characteristics of patients with OCD and controls recruited at Champalimaud Foundation. **Legend**: Results are presented in absolute number (%) or mean ± standard deviation. N – absolute number; OCD – obsessive-compulsive disorder, YBOCS – Yale Brown Obsessive-Compulsive Scale. * p-value < 0.05; ^1^Anholt et al. Psychol Med 2014; ^2^ According to Mini Neuropsychiatric Interview

|  | **Patients with OCD** | **Controls** | **p-value** |
| --- | --- | --- | --- |
| Number | 92 | 95 | - |
| Age, years | 36.00 ± 11.99 | 36.65 ± 12.07 | 0.69 |
| Sex, n (%) females | 51 (55.4) | 54 (56.8) | 0.88 |
| Education, years * | 13.91 ± 2.48 | 15.17 ± 2.58 | **0.001** |
| BMI, kg/m<sup>2</sup> | 24.81 ± 5.10 | 23.59 ± 3.29 | 0.057 |
| Smokers, n (%) | 23 (25.0) | 22 (23.2) | 0.86 |
| Age of onset, years | 16.6 ± 9.8 | - | - |
| Early-onset, n (%)<sup>1</sup> | 68 (73.9) | - | - |
| YBOCS-I, Total | 20.00 ± 6.32 | 0.56 ± 1.35 | < **0.0005** |
| Obsessions | 10.27 ± 3.25 | 0.38 ± 0.96 | < **0.0005** |
| Compulsions | 9.73 ± 3.82 | 0.22 ± 0.84 | < **0.0005** |
| STAI, Trait | 51.60 ± 13.92 | 30.88 ± 8.65 | < **0.0005** |
| State | 43.19 ± 14.00 | 28.22 ± 7.32 | < **0.0005** |
| BDI-II | 16.58 ± 10.96 | 5.09 ± 6.34 | < **0.0005** |
| Current depressive episode, n (%) <sup>2</sup> | 27 (29.3) | 0 (0) | < **0.0005** |
| Medicated, n (%) | 76 (84.4) | 1 (1.1) | < **0.0005** |
| Medication-naïve n (%) | 14 (15.6) | - | - |

A second sample of 47 patients with OCD and 36 healthy volunteers was recruited at University of Minho (UM; Braga, Portugal). Among these, age at inclusion, sex and years of education were similar in cases and controls. Patients had a mean age of 16.5±8.5 years at onset of OCD (early-onset subtype^24^ in 71.7%), with moderate to severe illness severity on average (total YBOCS-I 30.0±6.8)^25^ when recruited. Regarding pharmacological treatment, while almost 81% of patients were on psychotropic medication, no controls were on medication. Nine (19.1%) patients were medication-naïve. BMI, percentage of smokers, prevalence of active depressive episodes, and severity of anxiety and depressive symptoms were not assessed in this sample. When pooled the two samples had a total of 139 patients with OCD and 131 controls (**SupTable 1**).

### b. General medical conditions in patients with OCD and controls

When compared with healthy volunteers, patients recruited at the CF had a significantly higher prevalence of self-reported auto-immune disorders, allergies, asthma, irritable bowel syndrome, migraine and thyroid disorders (**Figure 2**). Comorbidity with general medical conditions was not assessed in the sample recruited at UM.

**Figure 2.**
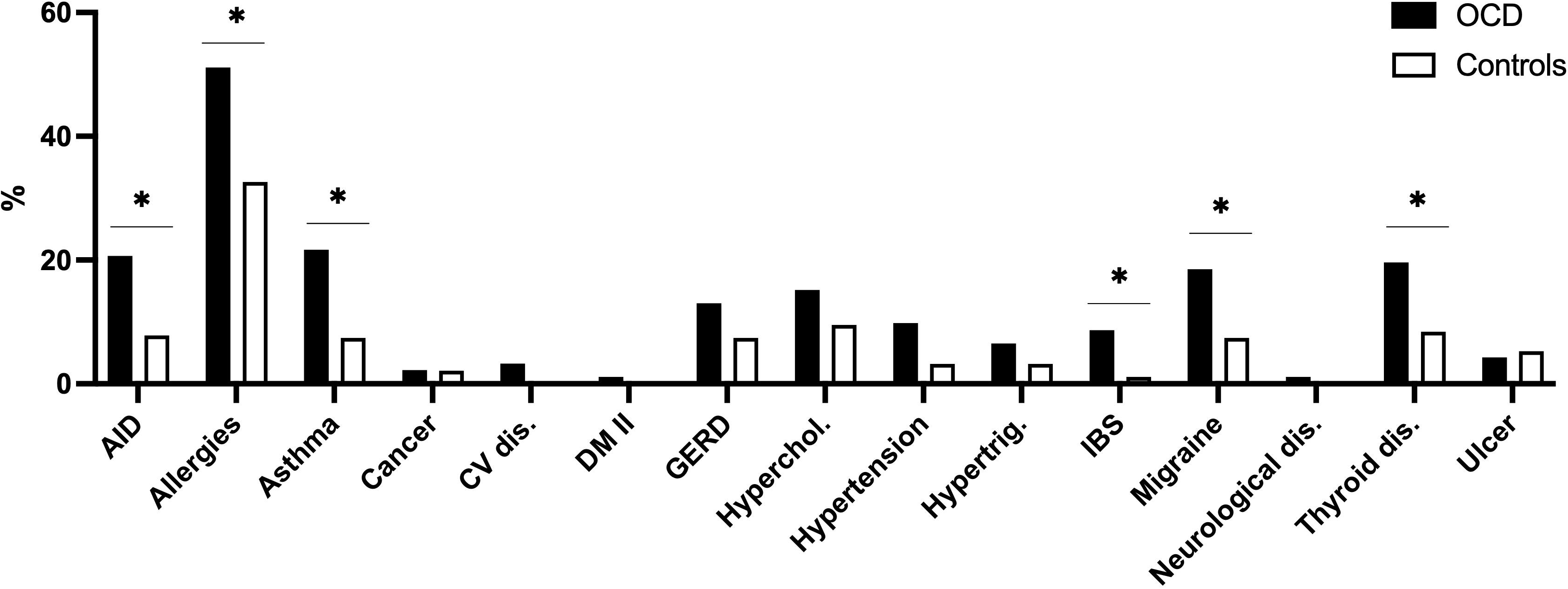
General medical conditions in patients with OCD and controls recruited at Champalimaud Foundation. **Legend:** AID - autoimmune disorders; CV - cardiovascular; DM II - diabetes mellitus e 2; GE -Gastroesophageal reflux diseases; Hyperchol - hipercholesterolemia; Hypertrig - Hipertriglyceridemia; IBS - irritable bowel syndrom; OCD - obsessive-compulsive disorder.

### c. Markers of immune dysfunction in patients with OCD and controls

In the sample recruited at CF, the mean concentration of high-sensitivity CRP (hsCRP) and the prevalence of individuals with high-hsCRP (i.e. ≥ 3 mg/dl^26^; **Figure 3**), as well as the peripheral concentration and gene expression of twelve cytokines (**Table 2, SupFigures 1-3**), was similar between groups. Similar results were found when comparing hsCRP and the concentration of the twelve cytokines between patients and controls recruited at UM, and when assessing both samples together (**Figure 3**, **Table 2)**.

**Figure 3.**
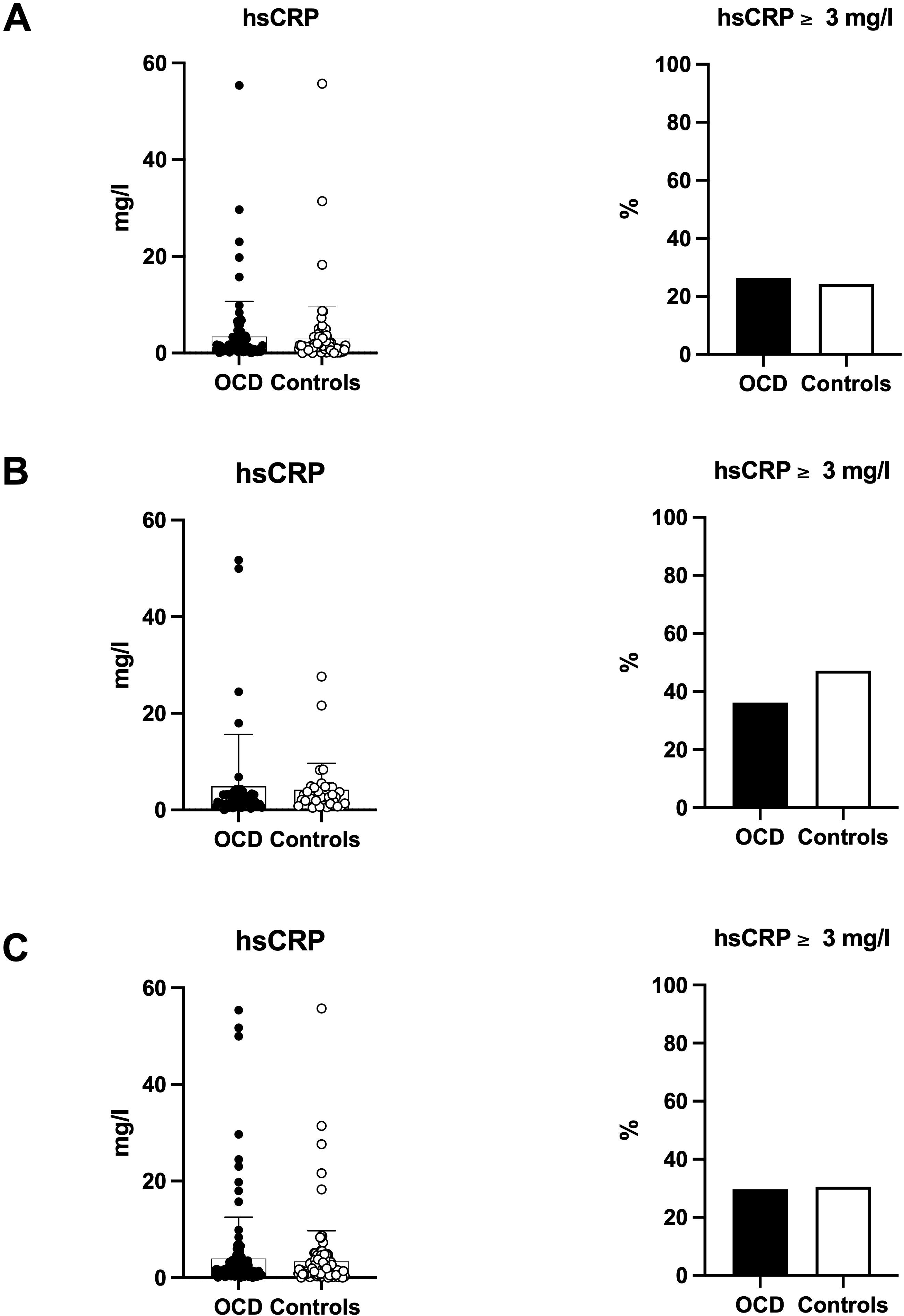
hsCRP concentration in patients with OCD and controls. A-Champalimaud Foundation; B - University of Minho; C - Champalimaud Foundation and University of Minho. **Legend:** hsCRP-high-sensitivity c-reactive protein; OCD-obsessive-compulsive disorder. Bars represent

**Table 2.**
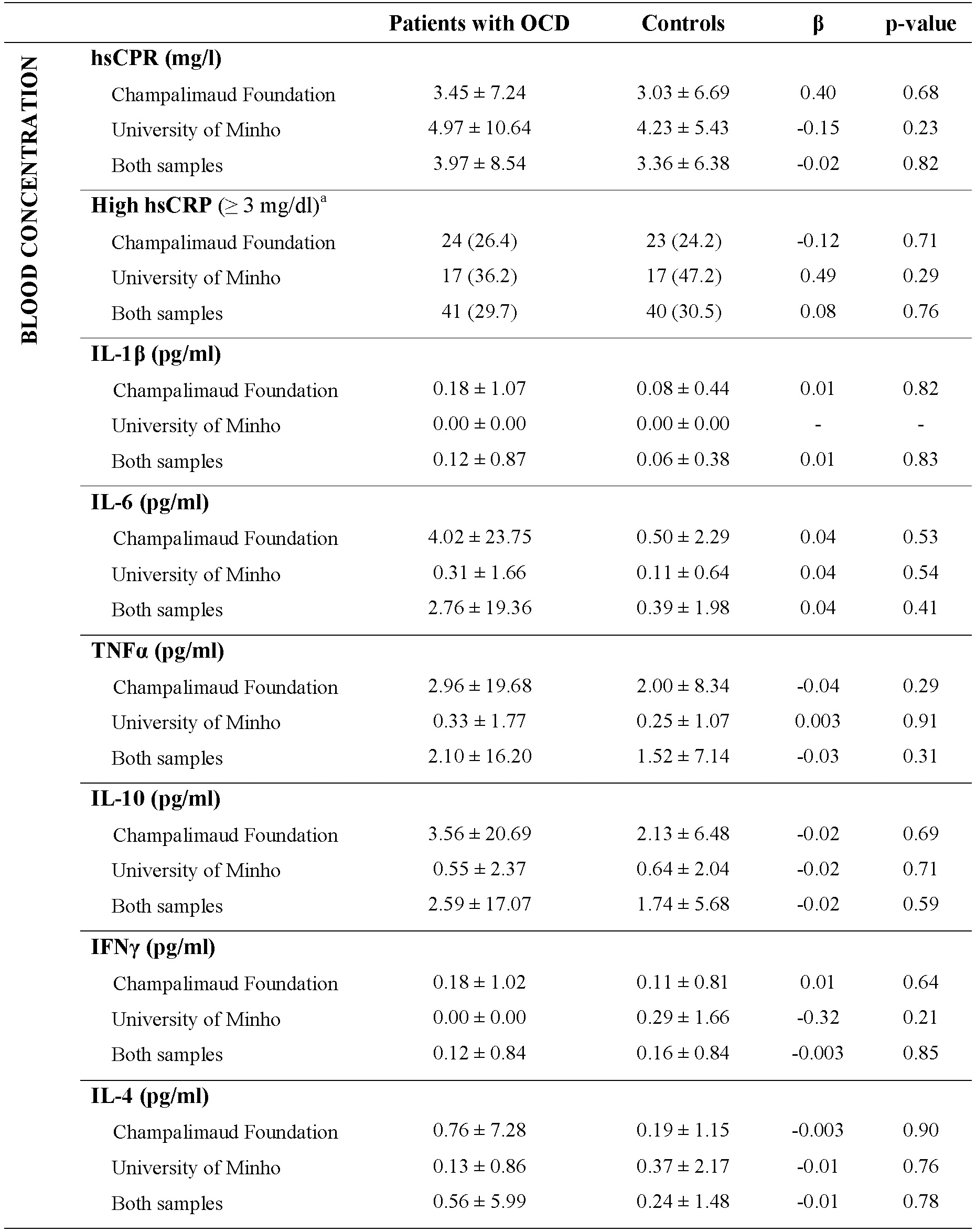
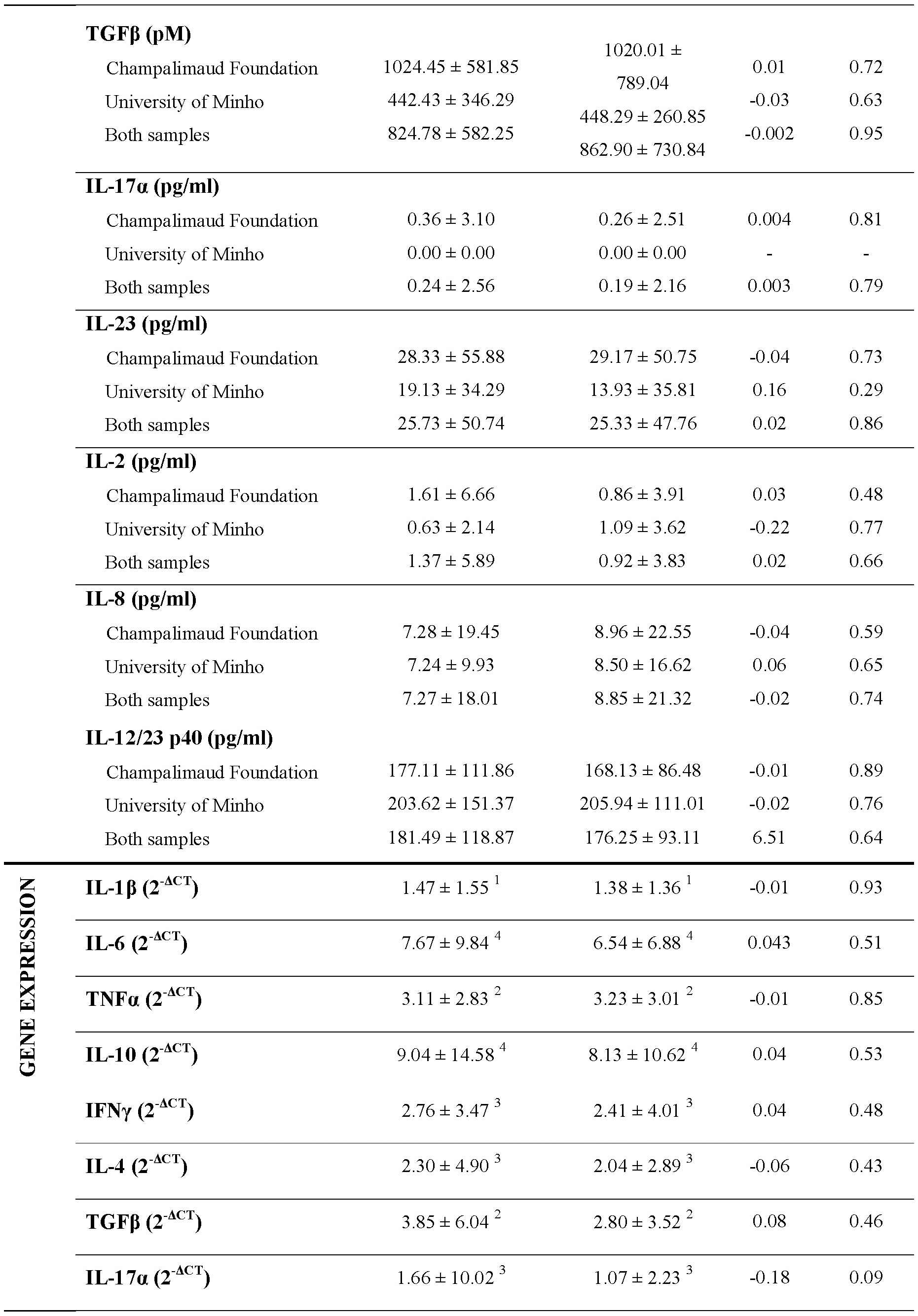
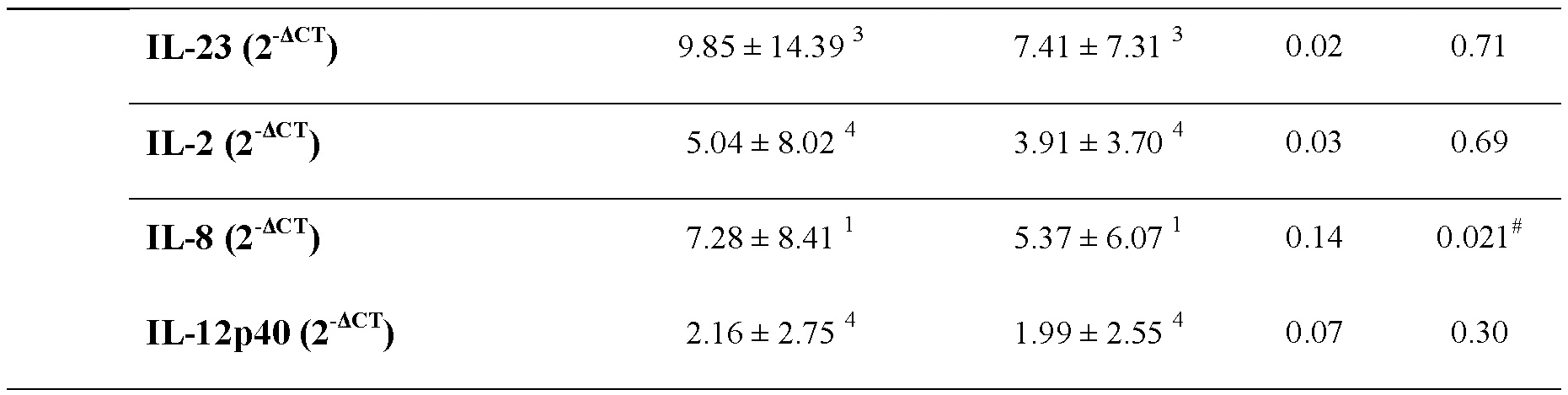
Markers of peripheral inflammation in patients with OCD and controls. **Legend**: Results are presented in mean ± SD or n _positives_ (%). Regressions for Champalimaud Foundation and University of Minho are adjusted for age and sex; regressions for both samples are adjusted for age, sex and center. The concentration of continuous variables were log-transformed for regression analyses. a. Odds ratio for high hsCRP in multiple logistic regressions: Champalimaud Foundation = 0.88; University of Minho = 1.63; Both samples = 1.09. 1. x10^-1;^ 2. X 10^-2^; 3. X 10^-3^; 4. X 10^-4^. β – unstandardized coefficient for the variable Group (i.e. OCD or control); IL – interleukin; IFN – interferon; OCD – obsessive-compulsive disorder; SD – standard deviation; TGF – transforming growth factor; TNF – tumor necrosis factor. ^#^ p-value did not remain significant after controlling for multiple testing using Benjamini-Hochberg’s procedure.

Subsequently, we compared the prevalence of anti-nuclear antibodies (ANAs), anti-thyroid peroxidase antibody (TP), anti-thyroglobulin antibody (TG) and ABGA across groups. Again, we did not find significant differences between cases and controls in any of the two samples. The prevalence for ANAs was higher in patients at a close to significant level in the sample recruited at CF (p=0.053). Furthermore, when samples from the two sites were analysed together, this effect was further supported (p=0.026), albeit statistical significance being lost upon correction for multiple comparisons (**Figure 4, SupTable 2)**.

**Figure 4.**
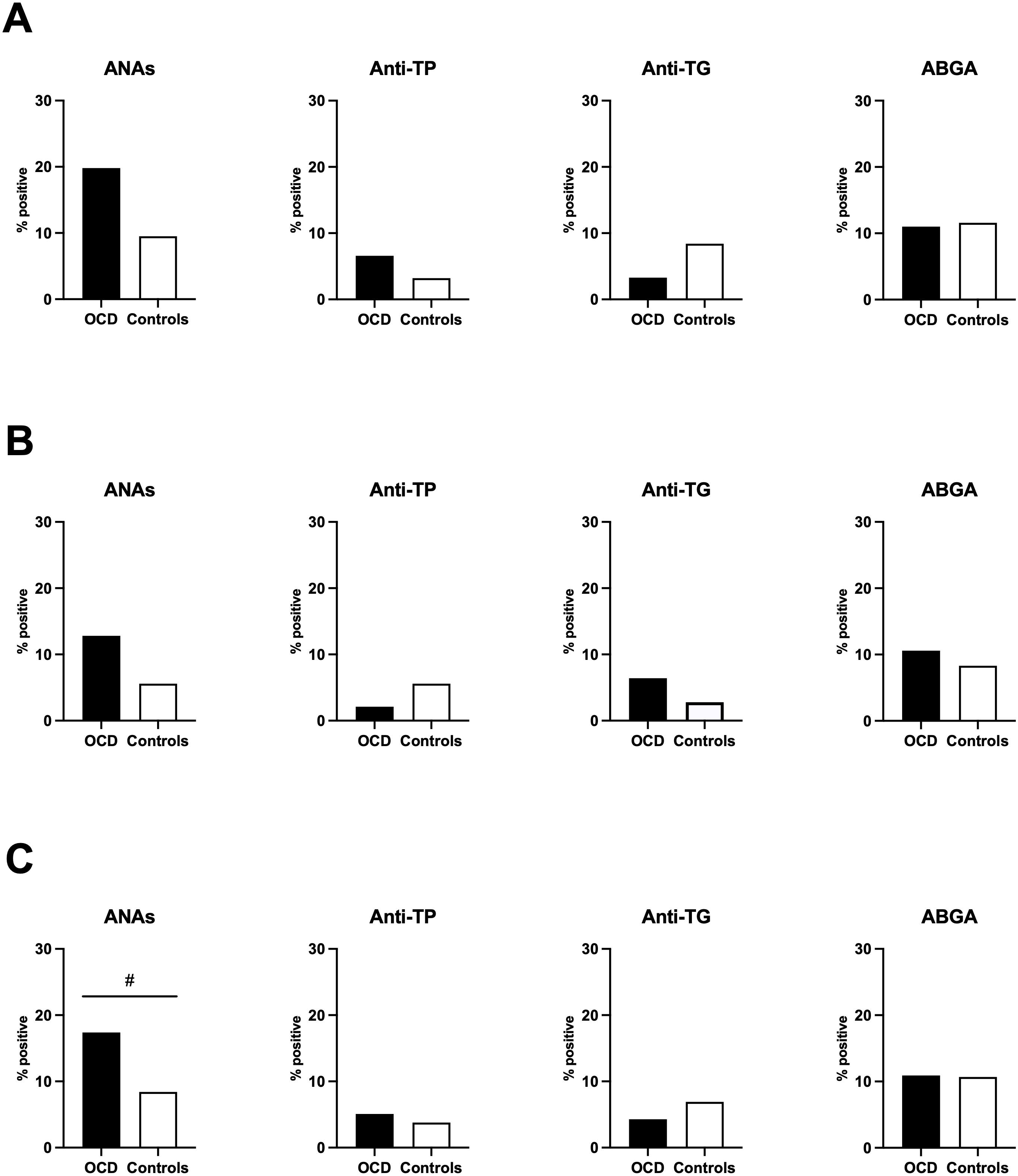
Prevalence of auto-antibodies in patients with OCD and controls. A - Champalimaud Foundation; B - University of Minho; C - Champalimaud Foundation and University of Minho. **Legend:** ABGA-Anti-basal ganglia antibodies; ANAs -Anti-nuclear antibodies; OCD - obsessive-compulsive disorder; TG - thyroglobulin; TP - thyroid peroxidase. # p-value < 0.05 but did not remain significant after controlling for multiple testing using Benjamini-Hochberg’s procedure.

### d. Subgroup analyses

To explore potential sources of heterogeneity, we performed subgroup analyses by comparing controls with specific patients’ subgroups. Patients with early-onset OCD had lower TNF-α concentration in the CF (p=0.02) and full (p=0.01) samples, and lower IL-17α gene expression in the CF sample (p=0.01), relative to controls. On the other hand, patients with late-onset OCD had lower hsCRP in the UM sample (p=0.02) and lower IL-12p40 gene expression in the CF sample (p=0.03). Patients without a current depressive episode, in the CF sample, had lower IL-17α gene expression relative to controls (p=0.01), while those who were naïve for psychotropic medication in the CF and full samples had higher IL-6 concentration (p<0.05 for both; **SupTable 3**). Prevalence for ANAs was higher in patients with early-onset OCD when compared to controls, only when assessing both samples together (**SupTable 4**). However, after correcting for multiple comparisons, none of these differences remained significant.

### e. Association with clinical characteristics among patients with OCD

Subsequently, we performed an exploratory analysis to assess if, among patients, immune markers were associated with clinical characteristics. Significant associations were not found for symptoms of anxiety nor medication status. Several immune markers were associated with age of onset, YBOCS-I, current depressive episode, antidepressant dose equivalent and history of autoimmune-disorder. In most cases these associations were only borderline significant and did not remain significant after correcting for multiple comparisons (**SupTable 5**). However, patients with early-onset OCD had significantly lower concentration of transforming growth factor (TGF)-β when compared to those with a late-onset of the disorder, even after correction (**SupFigure 4**).

## 4. Discussion

In this study, we tested the hypothesis that adults with OCD have a dysfunctional immune homeostasis and a chronic low-grade inflammatory state. We were inspired by the often cited, but poorly documented hypothesis that chronic inflammation plays a decisive role in the pathophysiology of OCD, similarly to what has been amply demonstrated in mood disorders and psychosis. To pursue this goal, we assessed a representative panel of markers of peripheral inflammation and general auto-antibodies in two samples of adult patients with OCD and controls, recruited in two different locations. We consistently found similar levels of systemic immune markers between groups, suggesting that patients with OCD do not have a significantly altered systemic immune profile, namely in cross-sectional assessments conducted in an adult symptomatic population.

We studied well known markers of peripheral inflammation, namely hsCRP and peripheral cytokines. hsCRP, an acute phase protein, is a validated indicator of acute and chronic ongoing inflammatory processes^27^, and a marker of subclinical low-grade peripheral inflammation in cardiovascular risk stratification (<1, 1 to 3 and >3 mg/L corresponding to low-, medium-, and high-risk categories, respectively)^28^. It has been widely studied in several neuropsychiatric disorders^29^, but to our knowledge to date only two studies have compared hsCRP between patients with OCD and healthy controls^30,31^. Although both found significantly higher concentrations of hsCRP in patients, the two studies had small samples sizes (21 and 33 patients) and one did not assess the impact of depressive symptoms in hsCRP levels^30^, which represents an important limitation given the fact that patients had significantly more depressive symptoms than controls, and that hsCRP has been demonstrated to be associated with depression^32^. In our study, we did not find differences in peripheral hsCRP between patients and controls, neither when assessing its concentration continuously, nor in analyses according to validated cut-offs. Furthermore, we assessed a total of 139 patients with OCD, making our study the largest publication to date evaluating hsCRP in outpatients with OCD, and with results remaining consistent across subgroup analyses, assessing patients without a current depressive episode or who were treatment-naïve, underlying the robustness of our findings.

Notwithstanding its high sensitivity, hsCRP is very unspecific^33^. We thus assessed the concentration and gene expression of twelve peripheral cytokines, that were specifically chosen for being representative of the four main T helper (Th) cells differentiation pathways [Th1, Th2, Th17 and T regulatory (Treg)] and of the two macrophage functional states (M1 and M2)^34–38^.

We did not find differences for any of the cytokines assessed, which is consistent with the similar hsCRP concentrations across groups. To date, two meta-analyses assessed peripheral cytokines in OCD, with conflicting results. The first focused specifically on three pro-inflammatory cytokines (i.e. IL-1β, IL-6 and TNF-α), and found a significant reduction of IL-1β in patients when compared to controls. However, this analysis included data from only four studies, including a total of 77 patients and 76 controls, which is approximately half of the sample size of our study^21^. The second meta-analysis included a higher number of studies and participants (sixteen studies including 538 patients and 463 controls), as well as a higher number of cytokines (i.e. IL-1β, IL-6 and TNF-α, IL-4, IL-10 and IFN-γ). Interestingly, no significant differences were found when comparing cytokines between groups^22^, which is in accordance with our findings. In secondary analyses, the first meta-analysis found significantly higher IL-6 in studies including adults without medication, and lower TNF-α in studies excluding individuals with comorbid depression^21^, while the second described higher IL-1β in studies assessing medication-naïve patients^22^. We consistently found non-significant results in subgroup analyses comparing patients who were medication-naïve or without a current depressive episode, with the exception of minor differences that did not survive correction for multiple comparisons. Furthermore, our results were consistent when assessing peripheral cytokines using two different approaches. First, we assessed the peripheral concentration of cytokines using ELISA, which is the most standard methodology in previous publications^21,22^. Subsequently, given that cytokines can be produced by several cells in addition to leukocytes, such as adipocytes, hepatocytes or endothelial cells^39^, we assessed cytokine gene expression in peripheral blood, which is restricted to cytokines produced by circulating blood cells. To our knowledge, this is the first study assessing cytokine gene expression in OCD, which considerably underlines the robustness of our negative findings.

To test whether auto-immune processes could represent an important mechanism in OCD pathophysiology, we also assessed three groups of autoantibodies, and consistently found similar prevalences in patients with OCD and controls. Although several studies have highlighted an increased prevalence of auto-immune and thyroid disorders in OCD^13–15^, to date only one study assessed ANAs and thyroid antibodies in this population, and found no significant differences in a group of anti-tissue antibodies, including ANAs, as well as in a group of anti-thyroid antibodies, including anti-TP and anti-TG^19^. However, it is worth mentioning that the control group in this study included patients with other psychiatric disorders, and to our knowledge, differences in these antibodies between patients with OCD and healthy controls had not yet been tested. Importantly, we found a trend towards significancy when comparing ANAs between patients and controls, suggesting that, in what concerns ANAs, absence of significant differences could result from low statistical power. However, in a systematic review and meta-analysis on inflammatory markers in psychiatric disorders, Yuan and colleagues underlined that the sample size required to detect differences in inflammatory markers between patients and controls could be relatively small (i.e. around 50 cases and 50 controls to achieve 80% power to detect a significant difference)^29^. In our study, we included a total of 139 patients and 131 controls and a significant difference between groups did not emerge, which supports that ongoing general autoimmunity may not seem be a distinctive feature in adults with symptomatic OCD.

In what concerns ABGA, Pearlman and colleagues published a meta-analysis suggesting that patients with OCD have a fivefold increased odds of ABGA seropositivity, when compared to controls. However, among the seven studies included, only three were individually significant^40–42^, namely a pediatric study assessing controls with streptococcal infections, neurological or autoimmune disorders^40^, a study comparing OCD with non-contemporaneous controls with psychiatric disorders^41^, and a meeting abstract that, as far as we are aware, has not been published^42^. Consequently, none of the studies that were individually significant was a case-control published study assessing adult patients with OCD and contemporaneous controls, which limits the generalizability of results.

In view of the present results, we propose that peripheral inflammation and auto-immune phenomena are not a distinctive characteristic of adult symptomatic patients with OCD and, if present, may require much larger sample sizes to be detected. Our negative results do not necessarily contradict the hypothesis of an immune-mediated OCD pathophysiology, since it is possible that inflammatory processes can be evident solely during immune activation or within the CNS, and/or play a role earlier in life, as a trigger but not as a perpetuating factor. Indeed, while mood and psychotic disorders tend to start in early adulthood^43^, most patients with OCD experience their first OC symptoms in childhood^24^, and it is reasonable to hypothesize that state-dependent inflammatory changes could only be detected in pediatric samples. It is also possible that inflammatory processes can be a perpetuating factor only in specific patient subgroups, which would be in accordance with our exploratory analyses suggesting that patients with an early-onset of OC symptoms have significantly lower concentrations of TGF-β, when compared to those with a late-onset of the disorder. TGF-β is a regulatory cytokine with important functions in immune homeostasis that has received attention as a potential biomarker in several neuropsychiatric disorders, such as depression or schizophrenia^44–46^. However, regarding OCD, to our knowledge only one study has assessed TGF-β in a sample of pediatric patients, limiting conclusions regarding the association between early-vs. late-onset subtypes and TGF-β concentration^47^. Early-onset OCD has been conceptualized as a more severe clinical subgroup^24^, and it is possible that our finding translates reduced regulatory immune function in patients with early-onset phenotype. However, immune markers were not significantly different between patients with early-onset OCD and controls in subgroup analyses. Furthermore, it is important to underline that cytokines are very unspecific and intrinsically interact with one another through mechanisms of pleiotropy, redundancy, synergism and antagonism^48^, with caution needed when drawing conclusions based on only one cytokine.

Our work should be interpreted in the light of potential limitations. First, BMI, smoking and comorbidity with general medical conditions, were not assessed in participants recruited at UM, and we were thus not able to control for the impact of these confounders in this second sample. Secondly, we had a relatively low number of patients who were medication-naïve, and secondary analyses assessing the impact of medication can be underpowered due to sample size. Furthermore, given the choice to assess a large panel of cytokines and of autoantibodies, the need to correct for multiple comparisons may have contributed to type II errors. However, when considering uncorrected statistical comparisons, consistent differences did not emerge across populations, comparisons or markers, nor within groups of markers with similar characteristics, suggesting low likelihood of false negative analyses. Lastly, although we assessed a representative panel of systemic immune markers, we did not assess immune markers in the CNS. Even though evidence suggests that there is reciprocal communication between central and peripheral immune processes, it remains unclear how peripheral immune markers reflect central immune dysfunction^48^. Notwithstanding the abovementioned limitations, our study stands out for its large sample size, and rigorous methodology, controlling for several important confounders and multiple comparisons. Furthermore, our negative findings were consistent when assessing cytokines using two different techniques, and when repeating the same analyses across two case-control samples, recruited in two distinct locations.

In conclusion, we found no evidence for a significant association between systemic immune markers and presence of OCD in adult symptomatic patients, underling the need to critically assess previous literature and to consider negative results when formulating hypothesis to guide upcoming research on OCD pathophysiology, towards the development of better clinical approaches and improved care.

## 4. Methods

The research protocol was conducted in accordance with the declaration of Helsinki for human studies of the World Medical Association and approved by the Ethics Committees of the three recruitment centres and of NOVA Medical School.

### a. Sample recruitment and clinical evaluation

Patients with a diagnosis of OCD according to DSM-5 criteria were recruited between October 2019 and May 2023 in three clinical centres in Lisbon, namely the Champalimaud Foundation, *Unidade Local de Saúde Lisboa Ocidental* and *Unidade Local de Saúde São José*. Adult symptomatic patients were first invited to participate by their attending psychiatrist, and those who accepted were then contacted by one of the researchers. Concomitantly, age and sex-matched control volunteers without previous or current history of psychiatric disorders were recruited from the general population in the same geographical area using standard recruitment procedures such as flyers and online postings. Interested respondents replied by email or telephone and were invited to participate via telephone. We performed matching according to sex and age within a range of ± 2 years. Patients and controls were included if they were between 18 and 65 years old, were fluent Portuguese speakers and did not meet any of the following exclusion criteria: any acute medical illness, history of substance abuse or dependence (other than nicotine) in the last 12 months, neurodevelopmental disorders, dementia or any form of cognitive impairment, active neurological disease, illiteracy or inability to understand the study’s instructions, and inability to provide informed consent.

At inclusion, after providing free and informed consent for participation in the study, participants were first interviewed by a psychiatrist or psychiatry trainee to assess for socio-demographic characteristics and psychopathology. The OCD module of the Structured Clinical Interview for DSM-5 was used to confirm OCD diagnosis with active symptoms, and the YBOCS-II to assess the severity of OC symptoms. The Mini International Neuropsychiatric Interview (MINI) was used to screen for psychiatric comorbidities, while symptoms of anxiety and depression were assessed using the State-Trait Anxiety Inventory (STAI) and the Beck Depression Inventory-II (BDI-II), respectively. All psychometric scales were in Portuguese and validated for the Portuguese population^49–51^.

Additionally, we assessed a second sample of patients with OCD and control volunteers that were recruited at UM (Braga, Portugal) between April 2019 and June 2023. Patients with an OCD diagnosis according to DSM-5 were identified at *Hospital de Braga* (Braga, Portugal), and controls with no history of psychiatric disorders and without current medication were identified from the general population in the same geographical area. Patients were assessed for OC symptom severity using the YBOCS-I, and were only included if they scored ≥ 16. To allow direct comparability between the two samples, YBOCS-II from the CF sample was converted to YBOCS-I by adapting individual scores to the magnitude of the scale^49^.

### b. Blood collection and immune markers selection

Venous blood samples of patients and controls were collected after clinical assessment. All samples were taken during the afternoon, between 4 and 5 pm, to control for diurnal variations of immune markers. Among the 187 included participants, only 1 patient did not collect blood due to needle phobia. Blood was centrifuged at site at 1200 rcf at room temperature for 10 minutes and serum was immediately collected and stored frozen at -80°C. Blood was collected and processed in the same day at Hospital de Braga (Braga, Portugal) or CF (Lisbon, Portugal). All laboratory experiments were performed by the same researchers (AM and SF) at CF. Systemic immune markers were chosen for being representative of diverse biological processes proposed to play a role in immune-mediated OCD pathophysiology. First, we assessed the concentration of a profile of markers of peripheral inflammation, namely hsCRP and twelve cytokines [IL-1β, IL-2 IL-4, IL-6, IL-8, IL-10, IL-12p40, IL-17α, IL-23, interferon (IFN)-γ, TGF-β and tumor necrosis factor (TNF)-α]. While hsCRP was chosen for being a sensitive, validated and widely used marker of peripheral inflammation, the twelve cytokines were specifically chosen for being representative of the four main T helper (Th) cells differentiation pathways and of the two macrophage functional states^34–38^. Subsequently, we assessed a group of auto-antibodies including eight ANAs [anti-double stranded deoxyribonucleic (dsDNA), anti-ribonucleoprotein (RNP), anti-Smith (Sm), anti-Ro [Sjögren’s Syndrome (SS)-A], anti-La (SS-B), anti-topoisomerase-I (Scl-70), anti-centromeric protein (CENP), and anti-histidyl-tRNA synthetase (Jo-1)], anti-TP, anti-TG and ABGA. While ANAs, anti-TP and anti-TG were selected for being increased in general medical conditions that have been previously associated with OCD^13–15^, ABGA was selected given the fact that autoimmune-mediated basal ganglia dysfunction was proposed as an important mechanism in the pathophysiology of immune-related OC symptoms, namely in PANDAS ^52^.

### c. Enzyme-linked immunosorbent assay (ELISA) procedure

The peripheral concentration of all immune markers was assessed using ELISA, with samples tested in duplicate, and with all plates having an equivalent number of patients and controls. The absorbance was read using an ELISA plate reader at 450 nm (Microplate Photometer HiPo MPP-96, Biosan SIA). All experiments and calculation of results were performed strictly according to the kit instructions. Whenever results were inconclusive, measurements were repeated, and to assure a conservative approach when assessing binary variables, samples with two successive inconclusive results were registered as negative. Regarding markers of peripheral inflammation, hsCRP was assessed in serum samples using the OriGene Human hsCRP ELISA kit (ref. EA101010) and cytokines were assessed in EDTA-plasma samples using Mabtech ELISA flex kits (refs. IL-1β 3416-1H-20; IL-2 3445-1H-20; IL-4 3410-1H-20; IL-6 3460-1H-20; IL-8 3560-1H-20; IL-10 3430-1H-20; IL-12/-23p40 3450-1H-20; IL-17α 3520-1H-6; IL-23 3457-1H-20; IFN-γ 3420-1H-20; TGF-β 3550-1H-20; and TNF-α 3512-1H-20). Regarding autoantibodies, while ANAs and ABGA were assessed in EDTA-plasma samples using Medipan | Generic assays ANAPro (ref. 4012) and AFG Bioscience Human ABGA Elisa (ref. EK714527) kits, respectively, anti-TP and anti-TG antibodies were assessed in heparin-plasma samples using Abcam Human Anti-Thyroid Peroxidase (ref. ab178632) and Human Anti-Thyroglobulin (ab178631) ELISA kits, respectively.

### d. Real-time quantitative polymerase chain reaction (RT-qPCR) procedure

Given that cytokines can be produced by several cells in addition to leukocytes, such as adipocytes, hepatocytes or endothelial cells ^39^, we assessed cytokine gene expression using RT-qPCR, which is restricted to cytokines produced by circulating blood cells. For this purpose, at CF a small amount of blood was immediately stored at -80°C with TRIzol^TM^ LS Reagent (Invitrogen). One patient and 2 controls had coagulated samples and so cytokine gene expression could not be assessed in these three participants. Total RNA was isolated from thawed samples as per manufacturer’s instructions. cDNA was synthetised using the QuantiTect® Reverse Transcription kit (Qiagen). RT-qPCR was performed in a QuantStudio™ 5 Real-Time PCR System, 384-well (Applied Biosystems™) using the SsoFast EvaGreen Supermix (Bio-Rad), in 10 μl reactions using 0,5 μl of each primer (10mM) (**SupTable 6)** and 1ul of the cDNA template.

All samples were run in triplicates. Cycling conditions included an enzyme activation step at 95°C for 30 seconds followed by 40 cycles of denaturation at 95°C for 5 seconds and of annealing/extension at 60°C for 5 seconds. This was followed by melting curve analysis that consisted of 1 cycle with an increase in temperature from 65°C to 95°C at 0.5°C/second. The amplified DNA was detected by fluorescence quantification of the double-stranded DNA binding dye EvaGreen and the relative quantification analysis was performed using the Design and Analysis Software v2.5.1. for QuantStudio 3 and 5 systems (Applied Biosystems™). To determine the level of gene expression, we tested three reference genes [glyceraldehyde 3-phosphate dehydrogenase (GAPDH), β2-microglobulin and β-actin] to control for experimental error. We tested between-group variability by comparing 2^-Cq^ between patients and controls^53^, intra-assay variability by comparing the coefficient of variation between the three triplicates and inter-assay variability by calculating the maximum fold change (MFC = minimum/maximum) and the coefficient of variation (%CV = SD/mean) for each reference gene ^54^. All reference genes had a non-significant variability between groups and a stable inter-assay expression (defined as a MFC lower than 2 and a mean expression lower than the maximum expression – 2 standard deviations) ^54^. However, GAPDH was the reference gene with lower inter-assay variability %CV (GADPH 6.95 %, β2-microglobulin 18.59%, and β-actin 7.39%) and was thus selected to normalize the mean value of the genes of interest. Four patients and 4 controls had a reference gene quantification cycle (C_q_) ≥ 2 standard deviations apart from the mean and were thus excluded from the analysis. ΔC_q_ was calculated by subtracting the C_q_ mean value of the three triplicates of GAPDH to the C_q_ mean value of the triplicates of the gene of interest. Next, 2^-ΔCq^ was calculated to account for the doubling of the target DNA during each performed RT-qPCR cycle. Whenever two or three of the triplicated C_q_s were found to have C_q_s > 40, 2^-ΔCT^ was assumed to be zero ^53,55^.

### e. Statistical analysis

When analyzing sociodemographic and clinical variables, comparison of categorical variables was performed using Fisher’s Exact test with odds ratio (OR) as a measure of effect size. For continuous variables, the independent samples t-test and the Welch t-test were used to compare the mean value of variables with and without homogeneity of variances, respectively, as assessed by Levene’s test for equality of variances.

To compare markers of systemic immune function between patients with OCD and control volunteers, consecutive multiple linear and logistic regressions were performed for each individual marker, analyzing the immune marker as dependent variable and group (OCD or control) as independent variable, adjusting for age at inclusion (in years) and sex (male or female). Given the fact that data for immune parameters are positively skewed due to their low concentrations, the concentration of hsCRP and cytokines were log-transformed^56,57^. hsCRP concentration was analyzed both as a continuous variable and categorized using validated cut-offs [i.e. high (≥ 3 mg/dl) vs. low (< 3 mg/dl) hsCRP ^26^. These specific cut-offs have well-known prognostic implications as markers of chronic low-grade inflammation in cardiovascular diseases [54]. Autoantibodies were assessed categorically by calculating positivity according to the cut-offs indicated in the kits’ instructions. The eight ANAs were assessed individually, and ANA positivity was considered if positivity was found for any of one of the ANAs. For each immune marker and autoantibody, to explore potential sources of heterogeneity that could be biasing results, we performed subgroup analyses by comparing controls with the following patients’ subgroups: early-onset OCD (≤19 years-old^24^), late-onset OCD (>19 years-old^24^), patients without a current depressive episode (according to Mini-Neuropsychiatric Interview), and those who were naïve for psychotropic medication. All abovementioned analyses were first performed in the sample recruited at CF, and subsequently in the sample recruited at UM. Lastly, to reduce bias associated with sample size, we assessed both samples together, also adjusting for site of recruitment (i.e. CF or UM).

Subsequently, we performed exploratory analyses in the pooled patient sample, to assess if immune markers were associated with clinical characteristics. To this aim, we performed sequential multiple linear and logistic regressions for each marker individually, assessing the immune marker as dependent variable and clinical characteristics [early-onset OCD^24^, severity of OC and anxiety symptoms, current depressive episode according to Mini-Neuropsychiatric Interview, medication status, antidepressant dose according to fluoxetine equivalent dosages^58^, and comorbidity with auto-immune disorders] as independent variables, adjusting for age at inclusion (in years), sex (male or female) and recruitment center (i.e. CF or UM). Given the exploratory character of these analyses and to ensure statistical power, patients from both samples were analyzed together.

We used the Benjamini-Hochberg’s procedure to control for multiple testing, assuming a 0.1 false discovery rate^59^. Given that the maximal proportion of missing data was less than 5% across all variables, missing data was addressed using listwise deletion. Data analysis was performed using SPSS (IBM Corp. Released 2015 SPSS Statistics for Windows, Version 23.0. Armonk, NK: IBM Corp) and GraphPad Prism (2022 GraphPad Prism for Windows, Version 9.4.1, San Diego, California USA) was used for the development of graphical content.

## Data Availability

All data produced in the present study are contained in the manuscript or available upon reasonable request to the authors.

## Acknowledgements

We would like to thank Diana Pereira MD, Sandra Nascimento MD, and Vânia Viveiros MD (*Unidade Local de Saúde São José)*, and Ana Sofia Sequeira MD and Catarina Santos MD (*Unidade Local de Saúde Lisboa Ocidental*) for their collaboration in participant recruitment, the technicians from the Champalimaud Clinical Center for their collaboration in blood collection, and the staff at the Champalimaud Foundation Biobank, Molecular and Transgenic Tools (MTT) Platform, and Champalimaud Glass Wash & Media Preparation Platform, for their availability and guidance in blood processing and analyses. We would also like to express our gratitude to all participants who generously accepted to contribute to scientific knowledge by taking part in our study.

## Authors contributions

AM, BBC, JO and AJOM conceived and designed the project. AM, ND, CF, TQ, PR, BBC, JO and AJOM developed and implemented the study at the CF, while PM, FM and ADC developed and implemented the study at UM. AM and PR were the Principal Investigators in the two external recruitment centers, namely *Unidade Local de Saúde Lisboa Ocidental* and *Unidade Local de Saúde São José*, respectively, and were responsible by the submission of the project to the respective Ethics Committees. AM, ND, CF, TQ, PM, ADC, BBC and JO collected data. AM, CF, ND and ADC elaborated and updated the database. AM and SF performed lab experiments. AM analyzed data, and wrote the original draft, with supervision and guidance from JO and AJOM. All authors had full access to anonymized data, performed data interpretation, revised the manuscript critically and approved the final version to be published.

