## Supplementary figures and images for "Immune (dys)function in obsessive-compulsive disorder"

### Supplementary Figure 1

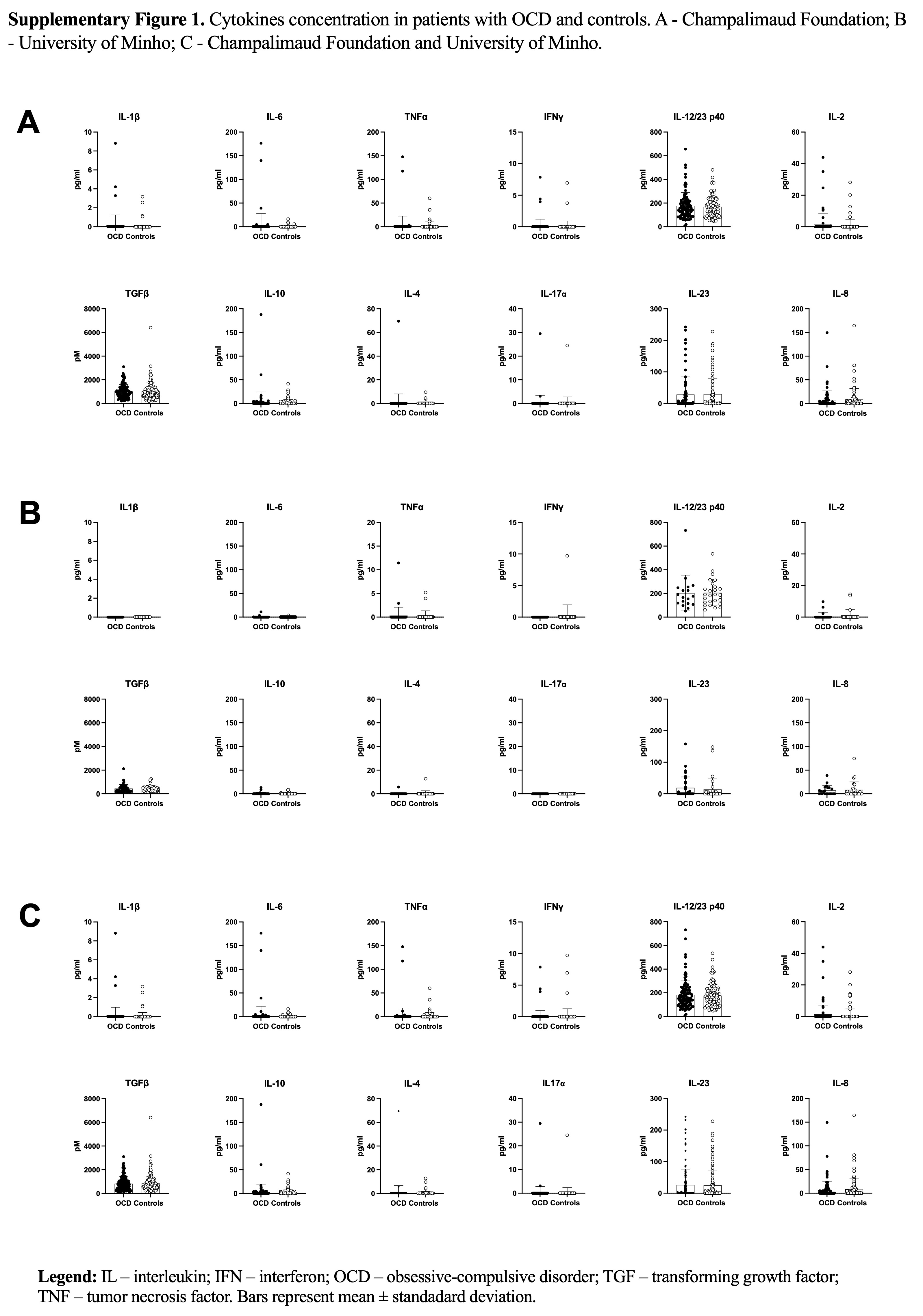

### Supplementary Figure 2

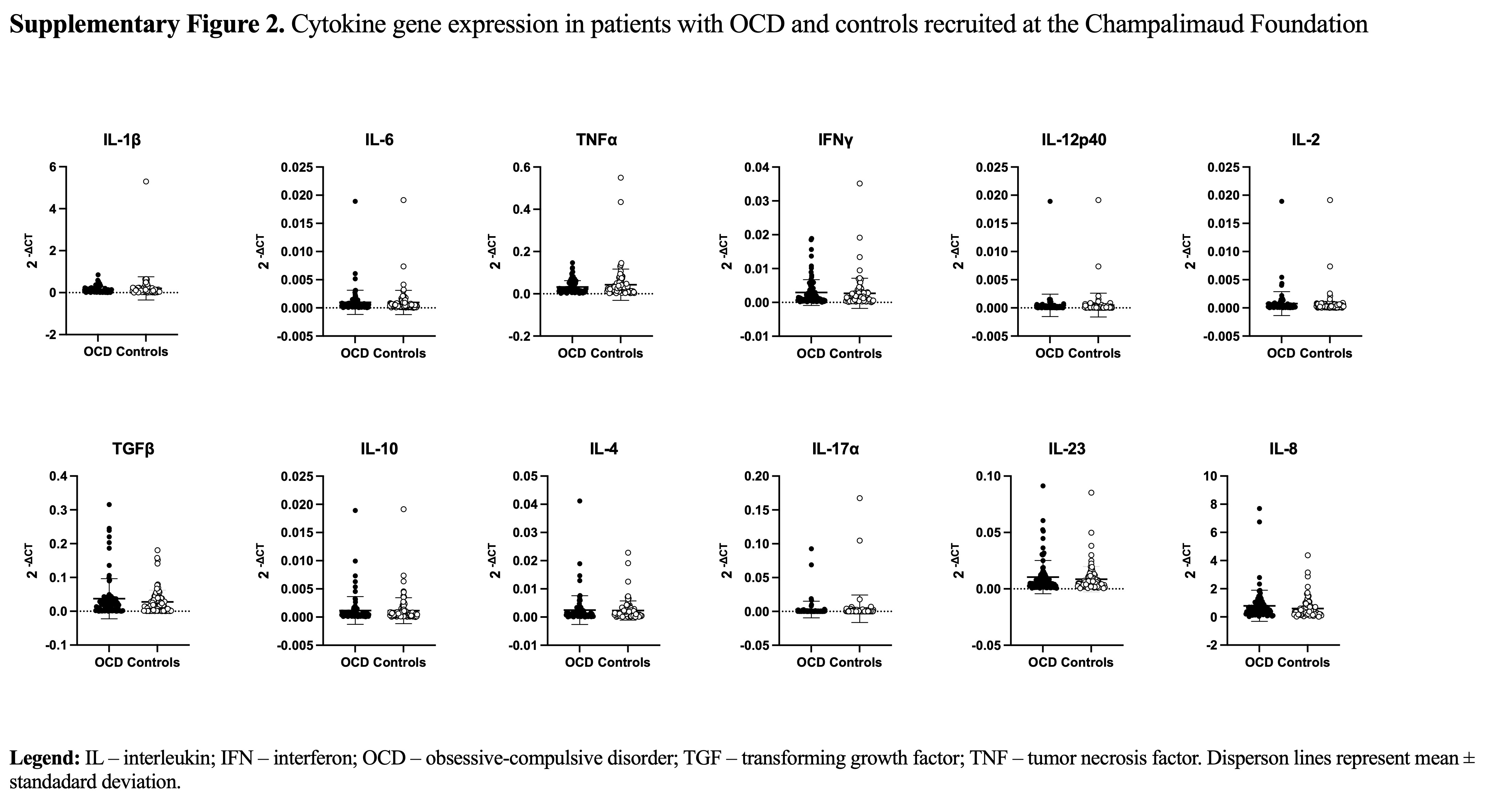

### Supplementary Figure 3

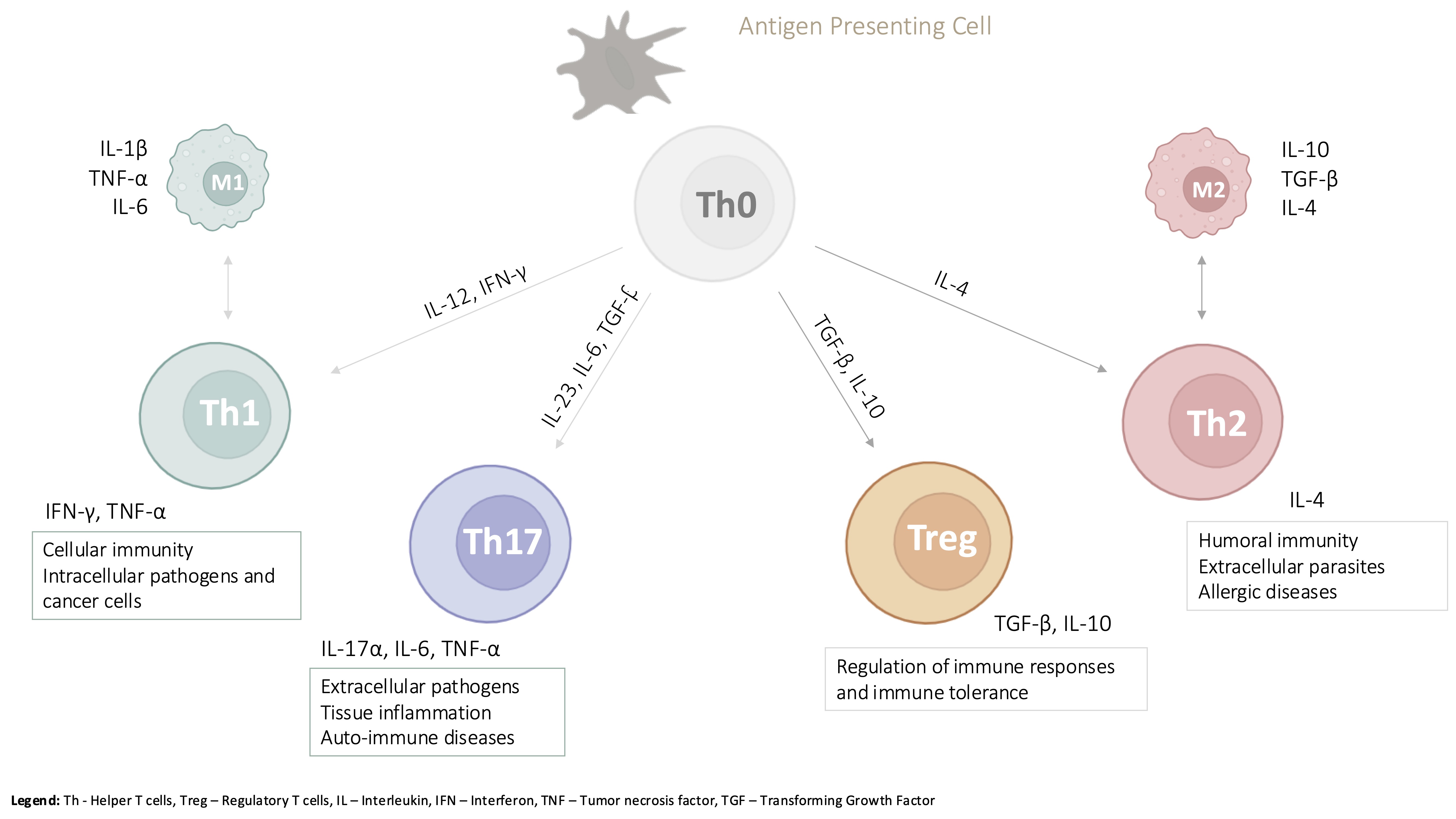

### Supplementary Figure 4

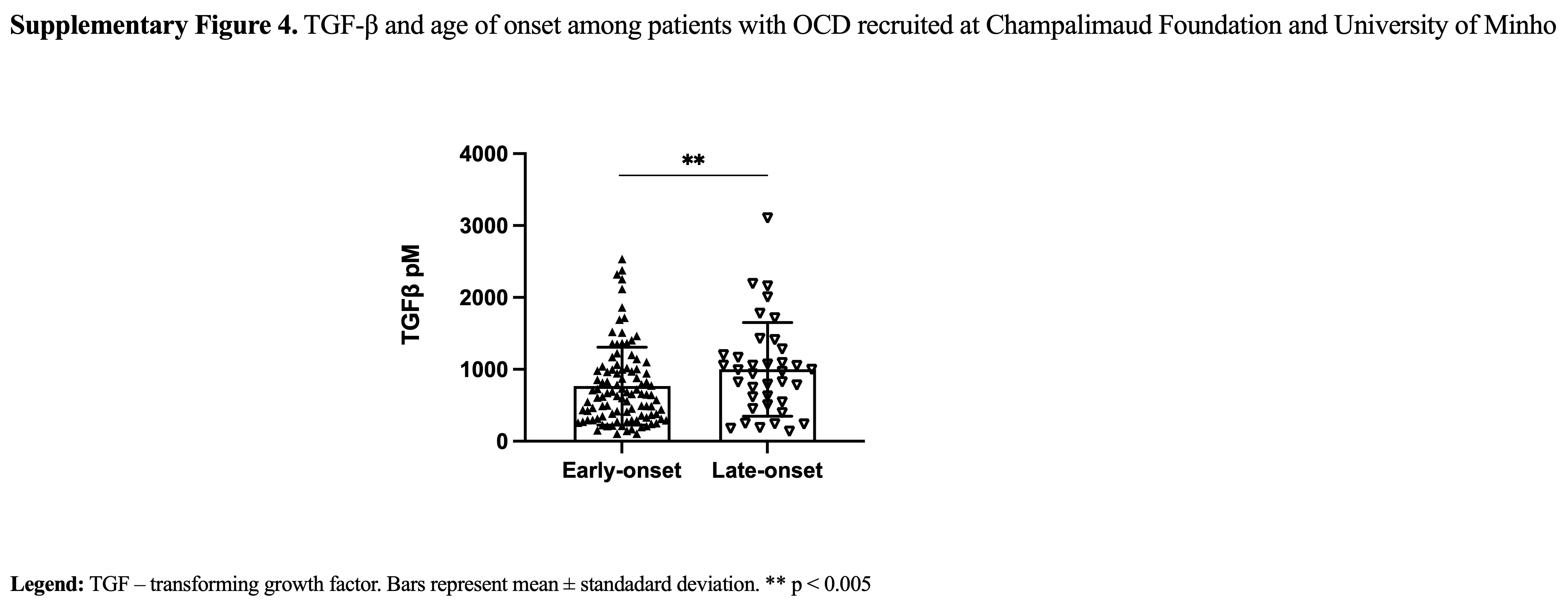
