## Supplementary Table 1 for "Immune (dys)function in obsessive-compulsive disorder"

**Supplementary Table 1** – Sociodemographic and clinical characteristics of patients with OCD and controls recruited at University of Minho and of both samples together

|  | **University of Minho** | | | **Champalimaud Foundation and University of Minho** | | |
| --- | --- | --- | --- | --- | --- | --- |
|  | **Patients with OCD** | **Controls** | **p-value** | **Patients with OCD** | **Controls** | **p-value** |
| Number, n | 47 | 36 | **-** | 139 | 131 | **-** |
| Age, years  Sex, n (%) females  Education, years | 28.91 ± 10.30  24 (51.1)  13.79 ± 5.58 | 29.78 ± 12.64  22 (47.8)  15.11 ± 3.48 | 0.73  0.38  0.10 | 33.58 ± 11.89  54  13.87 ± 2.89 | 34.76 ± 12.56  58  15.15 ± 2.84 | 0.43  0.54  **< 0.0005** |
| Age of onset, years  Early-onset, n (%) ^1^ | 16.51 ± 8.45  13 (71.7) | **-**  **-** | **-**  **-** | 16.58 ± 9.34  (73.2) | **-**  **-** | **-**  **-** |
| YBOCS-I, Total  Obsessions  Compulsions | 29.96 ± 6.67  14.41 ± 3.96  15.96 ± 2.62 | **-**  **-**  **-** | **-**  **-**  **-** | 23.22 ± 7.96  11.61 ± 3.98  11.74 ± 4.54 | **-**  **-**  **-** | **-**  **-**  **-** |
| Medicated, n (%)  Medication-naïve, n (%) | 38 (80.9)  9 (19.1) | 0 (0)  **-** | **< 0.0005**  **-** | 114 (83.2)  23 (16.8) | 1 (0.8)  **-** | **< 0.0005**  **-** |

**Legend**: Results are presented in absolute number (%) or mean ± standard deviation. N – absolute number; OCD – obsessive-compulsive disorder, YBOCS – Yale Brown Obsessive-Compulsive Scale. ^1^Anholt et al. Psychol Med 2014; ^2^ According to Mini Neuropsychiatric Interview.
