## Supplementary Table 2 for "Immune (dys)function in obsessive-compulsive disorder"

**Supplementary Table 2** – Auto-antibodies in patients with OCD and controls

|  | **Patients with OCD** | **Controls** | **OR** | **β** | **p-value** |
| --- | --- | --- | --- | --- | --- |
| **ANAs**  Champalimaud Foundation  University of Minho  Both samples | 18 (19.8)  6 (12.8)  24 (17.4) | 9 (9.5)  2 (5.6)  11 (8.4) | 0.43  0.36  0.42 | -0.85  -1.02  -0.87 | 0.053  0.23  0.026^#^ |
| **Anti-thyroid peroxidase**  Champalimaud Foundation  University of Minho  Both samples | 6 (6.6)  1 (2.1)  7 (5.1) | 3 (3.2)  2 (5.6)  5 (3.8) | 0.46  3.02  0.72 | -0.79  1.10  -0.33 | 0.28  0.38  0.59 |
| **Anti-thyroglobulin**  Champalimaud Foundation  University of Minho  Both samples | 3 (3.3)  3 (6.4)  6 (4.3) | 8 (8.4)  1 (2.8)  9 (6.9) | 2.70  0.34  1.52 | 0.99  -1.07  0.42 | 0.16  0.38  0.45 |
| **ABGA**  Champalimaud Foundation  University of Minho  Both samples | 10 (11.0)  5 (10.6)  15 (10.9) | 11 (11.6)  3 (8.3)  14 (10.7) | 1.08  0.63  0.96 | 0.07  -0.46  -0.04 | 0.87  0.57  0.92 |

**Legend**: Results are presented in mean ± SD or n _positives_ (%). ANAs – Anti-nuclear antibodies; OCD – obsessive-compulsive disorder. Regressions for Champalimaud Foundation and University of Minho are adjusted for age and sex; regressions for both samples are adjusted for age, sex and center. ^#^ p-value did not remain significant after controlling for multiple testing using Benjamini-Hochberg’s procedure.
