## Supplementary Table 3 for "Immune (dys)function in obsessive-compulsive disorder"

**Supplementary Table 3** – Markers of peripheral inflammation in patients’ subgroups and controls

|  |  | **Controls** | **Patients with EO-OCD** | **p-value** | **Patients with LO-OCD** | **p-value** | **Patients without depression** | **p-value** | **Medication-naïve patients** | **p-value** |
| --- | --- | --- | --- | --- | --- | --- | --- | --- | --- | --- |
| **BLOOD CONCENTRATION** | **hsCRP (mg/l)**  Champ. Foundation  University of Minho  Both samples | 3.03±6.7  4.23±5.43  3.36±6.38 | 3.23±5.28  6.18±12.51  4.2±8.43 | 0.49  0.62  0.67 | 4.06±11.16  1.98±1.78  3.33±9.04 | 0.81  **0.02#**  0.17 | 3±5.42  -  - | 0.95  -  - | 3.12±6.19  5,14±7,34  3,12±6,19 | 0.92  0.29  0.59 |
|  | **High hsCRP (≥ 3 mg/dl)**  Champ. Foundation  University of Minho  Both samples | 23 (24.2)  17 (47.2)  40 (30.5) | 19 (28.4)  13 (39.4)  32 (32.0) | 0.50  0.44  0.92 | 5 (20.8)  3 (23.1)  8 (21.6) | 0.68  0.17  0.25 | 15 (23.4)  -  - | 0.94  -  - | 3 (23.1)  6 (66.7)  9 (40.9) | 0.94  0.35  0.51 |
|  | **IL-1β (pg/ml)**  Champ. Foundation  University of Minho  Both samples | 0.08±0.44  0.00±0.00  0.06±0.38 | 0.19±1.19  0.00±0.00  0.13±0.97 | 0.81  -  0.80 | 0.14±0.67  0.00±0.00  0.09±0.54 | 1.00  -  1.00 | 0.2±1.21  -  - | 0.80  -  - | 0±0  0±0  0±0 | 0.42  -  0.52 |
|  | **IL-6 (pg/ml)**  Champ. Foundation  University of Minho  Both samples | 0.5±2.29  0.11±0.64  0.39±1.98 | 4.87±27.28  0.45±1.98  3.41±22.4 | 0.38  0.44  0.26 | 1.64±8.01  0.00±0.00  1.06±6.45 | 0.87  0.44  0.73 | 5.71±28.22  -  - | 0.19  -  - | 13.77±39.39  0,37±1,12  6,76±27,64 | **0.045#**  0.55  **0.043#** |
|  | **TNF-α (pg/ml)**  Champ. Foundation  University of Minho  Both samples | 2±8.34  0.25±1.07  1.52±7.14 | 0.0±0.0  0.0±0.0  0.0±0.0 | **0.02#**  0.16  **0.01#** | 11.23±37.66  1.1±3.21  7.67±30.55 | 0.44  0.26  0.31 | 2.37±18.47  -  - | 0.28  -  - | 0.28±0.99  1,27±3,81  0,28±0,99 | 0.44  0.31  0.75 |
|  | **IL-10 (pg/ml)**  Champ. Foundation  University of Minho  Both samples | 2.13±6.48  0.64±2.04  1.74±5.68 | 4.6±24.02  0.08±0.42  3.2±20.02 | 0.80  0.10  0.84 | 0.68±2.8  1.69±4.2  1.04±3.33 | 0.07  0.34  0.27 | 3.62±23.5  -  - | 0.52  -  - | 1.04±3.75  1,45±4,34  1,04±3,75 | 0.35  0.89  0.52 |
|  | **IFN-γ (pg/ml)**  Champ. Foundation  University of Minho  Both samples | 0.11±0.81  0.29±1.66  0.16±1.09 | 0.18±1.07  0.0±0.0  0.12±0.89 | 0.87  0.32  0.68 | 0.18±0.9  0.0±0.0  0.12±0.72 | 0.54  0.50  1.00 | 0.25±1.21  -  - | 0.42  -  - | 0.0±0.0  0.0±0.0  0.0±0.0 | 0.65  0.49  0.38 |
|  | **IL-4 (pg/ml)**  Champ. Foundation University of Minho  Both samples | 0.19±1.15  0.37±2.17  0.24±1.48 | 0±0.0±0.0  0±0.0±0.0  0±0.0±0.0 | 0.14  0.34  0.07 | 2.89±14.18  0.44±1.58  2.03±11.43 | 0.22  0.65  0.22 | 1.09±8.68  -  - | 0.64  -  - | 0.0±0.0  0,63±1,9  0.0±0.0 | 0.57  0.59  0.99 |
|  | **TGF β (pM)**  Champ. Foundation  University of Minho  Both samples | 1020.01±789.04  448.29±260.85  862.9±730.84 | 926.83±545.59  434.66±365.79  764.41±543.87 | 0.39  0.50  0.25 | 1308.85±602.49  453.98±318.79  1000.14±660.63 | **0.01#**  0.93  **0.045#** | 998.88±591.62  -  - | 0.97  -  - | 1070.82±385.3  559,09±267,39  1070,82±385,3 | 0.29  0.55  0.27 |
|  | **IL-17α (pg/ml)**  Champ. Foundation  University of Minho  Both samples | 0.26±2.51  0±0.0±0.0  0.19±2.16 | 0.05±0.38  0.0±0.0  0.03±0.32 | 0.63  -  0.64 | 1.23±6.02  0±0.0±0.0  0.8±4.84 | 0.30  -  0.29 | 0.51±3.7  -  - | 0.57  -  - | 0.0±0.0  0.0±0.0  0.0±0.0 | 0.68  -  0.76 |
|  | **IL-23 (pg/ml)**  Champ. Foundation  University of Minho  Both samples | 29.17±50.75  13.93±35.81  25.33±47.76 | 28.28±55.71  17.26±35.98  25.47±51.42 | 0.82  0.63  0.99 | 28.48±57.56  22.61±33.64  26.52±50.41 | 0.83  0.24  0.69 | 28.21±54.68  -  - | 0.96  -  - | 30.72±60.7  8,97±18,53  30,72±60,7 | 0.78  0.68  0.66 |
|  | **IL-2 (pg/ml)**  Champ. Foundation  University of Minho  Both samples | 0.86±3.92  1.09±3.62  0.92±3.83 | 1.35±5.52  0.89±2.66  1.25±5.03 | 0.61  1.00  0.70 | 2.33±9.21  0.23±0.73  1.71±7.76 | 0.42  0.58  0.68 | 2.27±7.82  -  - | 0.16  -  - | 1.36±3.55  2,01±3,9  1,36±3,55 | 0.35  0.58  0.29 |
|  | **IL-8 (pg/ml)**  Champ. Foundation  University of Minho  Both samples | 8.96±22.55  8.5±16.63  8.85±21.32 | 5.45±9.71  4.94±5.97  5.36±9.14 | 0.68  1.00  0.66 | 12.39±34.24  7.38±9.59  11.39±30.82 | 0.48  0.72  0.64 | 8.66±22.27  -  - | 0.99  -  - | 9.93±21.53  10,87±18,57  9,93±21,53 | 0.88  0.83  0.92 |
|  | **IL-12/23 p40 (pg/ml)**  Champ. Foundation  University of Minho  Both samples | 168.13±86.48  205.94±111.01  176.25±93.11 | 189.02±121.87  230.72±181.09  194.9±131.16 | 0.93  0.89  0.89 | 143.87±69.1  161.04±82.49  147.75±71.24 | 0.25  0.29  0.12 | 164.81±97.98  -  - | 0.59  -  - | 193.64±146.96  461,3±382,83  193,64±146,96 | 0.43  0.20  0.21 |
| **GENE EXPRESSION** | **IL-1β (2^-ΔCT^)** ^1^ | 1,39±1,36 | 1,27±1,32 | 0.37 | 2,06±1,98 | 0.27 | 1,25±1,22 | 0.55 | 1,25±1,17 | 0.55 |
|  | **IL-6 (2^-ΔCT^)** ^4^ | 6,54±6,88 | 7,35±8,78 | 0.69 | 8,63±12,65 | 0.6 | 6,6±7,94 | 0.9 | 13±17,76 | 0.13 |
|  | **TNFα (2^-ΔCT^)** ^2^ | 3,23±3,01 | 2,69±2,21 | 0.40 | 4,31±3,97 | 0.3 | 2,74±2,57 | 0.42 | 3,24±2,03 | 0.64 |
|  | **IL-10 (2^-ΔCT^)** ^4^ | 8,13±10,62 | 8,64±15,24 | 0.99 | 10,2±12,74 | 0.27 | 8,37±14,18 | 0.75 | 12,28±17,03 | 0.43 |
|  | **IFNγ (2^-ΔCT^)** ^3^ | 2,41±4,01 | 3,1±3,76 | 0.27 | 1,79±2,28 | 0.21 | 2,5±3,14 | 0.79 | 1,78±3,24 | 0.07 |
|  | **IL-4 (2^-ΔCT^)** ^3^ | 2,04±2,89 | 2,69±5,6 | 0.71 | 1,19±1,17 | 0.12 | 1,77±2,35 | 0.32 | 1,24±1,17 | 0.27 |
|  | **TGFβ (2^-ΔCT^)** ^2^ | 2,8±3,52 | 4,01±6,25 | 0.25 | 3,38±5,53 | 0.59 | 3,03±4,94 | 0.93 | 1,58±2,86 | 0.07 |
|  | **IL-17α (2^-ΔCT^)** ^3^ | 1,07±2,24 | 0,5±1,31 | **0.01#** | 5,05±19,63 | 0.39 | 0,49±1,15 | **0.01#** | 0,31±0,3 | 0.38 |
|  | **IL-23 (2^-ΔCT^)** ^3^ | 7,42±7,31 | 8,21±9,63 | 0.97 | 14,62±23 | 0.52 | 7,11±8,47 | 0.51 | 8,49±11,03 | 0.86 |
|  | **IL-2 (2^-ΔCT^)** ^4^ | 3,91±3,71 | 4,95±6,69 | 0.58 | 5,32±11,23 | 0.6 | 4,26±6,22 | 0.89 | 2,16±1,64 | 0.16 |
|  | **IL-8 (2^-ΔCT^)** ^1^ | 5,37±6,07 | 6,9±5,73 | **0.048#** | 8,4±13,65 | 0.09 | 5,87±4,49 | 0.21 | 3,83±3,15 | 0.27 |
|  | **IL-12p40 (2^-ΔCT^)** ^4^ | 1,99±2,55 | 1,94±2,82 | 0.91 | 2,81±2,51 | **0.03#** | 2,02±1,98 | 0.18 | 2,28±2,25 | 0.37 |

**Legend**: Results are presented in mean ± SD or n _positives_ (%). Regressions for Champalimaud Foundation and University of Minho are adjusted for age and sex; regressions for both samples are adjusted for age. sex and center. The concentrations of continuous variables were log-transformed for regression analyses. 1. x10^-1;^ 2. X 10^-2^; 3. X 10^-3^; 4. X 10^-4^. β – unstandardized coefficient for the variable Group (i.e. OCD or control); IL – interleukin; IFN – interferon; OCD – obsessive-compulsive disorder; SD – standard deviation; TGF – transforming growth factor; TNF – tumor necrosis factor. ^#^ p-value did not remain significant after controlling for multiple testing using Benjamini-Hochberg’s procedure.
