## Supplementary Table 4 for "Immune (dys)function in obsessive-compulsive disorder"

**Supplementary Table 4** – Auto-antibodies in patients’ subgroups and controls

|  | **Controls** | **Patients with EO-OCD** | **p-value** | **Patients with LO-OCD** | **p-value** | **Patients without depression** | **p-value** | **Medication-naïve patients** | **p-value** |
| --- | --- | --- | --- | --- | --- | --- | --- | --- | --- |
| **ANAs**  Champalimaud Foundation  University of Minho  Both samples | 9 (9.5)  2 (5.6)  11 (8.4) | 15 (22.4)  5 (15.2)  20 (20.0) | 0.39  0.15  **0.02#** | 3 (12.5)  1 (7.7)  4 (10.8) | 0.44  0.66  0.40 | 16 (25.0)  -  - | 0.98  -  - | 3 (23.1)  1 (7.1)  4 (14.8) | 0.11  0.69  0.17 |
| **Anti-thyroid peroxidase**  Champalimaud Foundation  University of Minho  Both samples | 3 (3.2)  2 (5.6)  5 (3.8) | 4 (6.0)  1 (3.0)  5 (5.0) | 0.40  0.55  0.68 | 2 (8.3)  0 (0)  2 (5.4) | 0.13  1.00  0.38 | 3 (4.7)  -  - | 0.73  -  - | 1 (7.7)  1 (7.1)  2 (7.4) | 0.29  0.91  0.44 |
| **Anti-thyroglobulin**  Champalimaud Foundation  University of Minho  Both samples | 8 (8.4)  1 (2.8)  9 (6.9) | 2 (3.0)  3 (9.1)  5 (5.0) | 0.22  0.16  0.78 | 1 (4.2)  0 (0)  1 (2.7) | 0.33  1.00  0.28 | 3 (4.7)  -  - | 0.43  -  - | 0 (0)  1 (7.1)  1 (3.7) | 1.00  0.38  0.59 |
| **ABGA**  Champalimaud Foundation  University of Minho  Both samples | 11 (11.6)  3 (8.3)  14 (10.7) | 5 (7.5)  5 (15.2)  10 (10.0) | 0.30  0.17  0.86 | 5 (20.8)  0 (0)  5 (13.5) | 0.07  1.00  0.40 | 9 (14.1)  -  - | 0.71  -  - | 0 (0)  1 (7.1)  1 (3.7) | 1.00  0.79  0.31 |

**Legend**: Results are presented in mean ± SD or n _positives_ (%). ANAs – Anti-nuclear antibodies; EO – early-onset; LO – late-onset; OCD – obsessive-compulsive disorder. Regressions for Champalimaud Foundation and University of Minho are adjusted for age and sex; regressions for both samples are adjusted for age, sex and center. ^#^ p-value did not remain significant after controlling for multiple testing using Benjamini-Hochberg’s procedure.
