## Supplementary Table 5 for "Immune (dys)function in obsessive-compulsive disorder"

**Supplementary Table 5** – Association of systemic immune markers and clinical characteristics in patients with OCD recruited at the Champalimaud Foundation and University of Minho

| **Patients with OCD** | | **Early-onset ^1^** | **YBOCS-I** | **STAI-T** | **STAI-S** | **Dep. Episode** | **Med. status** | **AD dose eq.** | **AID** |
| --- | --- | --- | --- | --- | --- | --- | --- | --- | --- |
| **hsCRP**  **(mg/dl)** | **β**  **p-val** | -0.25  **0.048#** | 0.001  0.86 | 0.002  0.73 | 4.99x10^-5^  0.99 | 0.14  0.30 | 0.04  0.76 | 0.003  **0.039#** | 0.20  0.21 |
| **IL-1β**  **(pg/ml)** | **β**  **p-val** | 0.004  0.93 | 0.00  0.91 | -0.002  0.39 | -0.001  0.52 | -0.001  0.99 | 0.03  0.52 | 0.00  0.46 | -0.05  0.45 |
| **IL-6**  **(pg/ml)** | **β**  **p-val** | -0.12  0.24 | -0.01  0.20 | -0.01  0.21 | -0.01  0.24 | -0.23  0.06 | -0.19  0.08 | 0.00  0.88 | 0.06  0.65 |
| **TNFα**  **(pg/ml)** | **β**  **p-val** | 0.11  **0.023#** | -0.004  0.17 | 0.00  0.94 | 0.00  0.95 | 0.01  0.89 | -0.01  0.87 | 0.00  0.60 | 0.07  0.37 |
| **IL-10**  **(pg/ml)** | **β**  **p-val** | -0.04  0.59 | 0.001  0.77 | 0.001  0.80 | 0.003  0.29 | 0.08  0.40 | 0.02  0.79 | -0.001  0.23 | 0.21  **0.045^#^** |
| **IFNγ**  **(pg/ml)** | **β**  **p-val** | 0.02  0.41 | 0.00  0.76 | 0.00  0.79 | 0.00  0.73 | -0.03  0.34 | 0.02  0.55 | -6.79x10^-5^  0.80 | -0.03  0.36 |
| **IL-4**  **(pg/ml)** | **β**  **p-val** | 0.06  0.08 | -0.003  0.24 | -0.002  0.30 | -0.002  0.29 | -0.03  0.46 | -0.01  0.80 | 0.00  0.60 | -0.04  0.46 |
| **TGFβ**  **(pM)** | **β**  **p-val** | 0.16  **0.003** | -0.002  0.59 | -7.64x10^-5^  0.97 | 0.00  0.81 | 0.053  0.38 | -0.09  0.14 | -0.001  0.20 | 0.11  0.10 |
| **IL-17α**  **(pg/ml)** | **β**  **p-val** | 0.03  0.16 | -0.002  0.16 | -0.001  0.34 | -0.001  0.35 | -0.03  0.39 | 0.01  0.64 | 2.90x10^-6^  0.99 | -0.02  0.66 |
| **IL-23**  **(pg/ml)** | **β**  **p-val** | 0.05  0.74 | -0.02  0.11 | -0.01  0.19 | -0.01  0.09 | -0.11  0.49 | 0.19  0.25 | 0.003  0.08 | -0.27  0.14 |
| **IL-2**  **(pg/ml)** | **β**  **p-val** | -0.03  0.70 | -0.01  **0.047#** | -0.004  0.17 | -0.003  0.26 | -0.17  **0.036#** | -0.08  0.34 | -0.001  0.45 | -0.04  0.67 |
| **IL-8**  **(pg/ml)** | **β**  **p-val** | -0.02  0.89 | -0.01  0.16 | -0.004  0.31 | -0.002  0.61 | -0.12  0.28 | 0.01  0.95 | -0.001  0.55 | 0.01  0.91 |
| **IL-12/23p40**  **(pg/ml)** | **β**  **p-val** | -0.05  0.42 | -0.01  0.18 | -0.001  0.80 | -0.001  0.55 | 0.06  0.33 | -0.11  0.14 | -0.001  0.09 | 0.03  0.67 |
| **ANAs**  **(% _positives_)** | **β**  **p-val** | 0.72  0.26 | -0.08  **0.036#** | -0.04  0.08 | -0.04  0.14 | 1.41  0.08 | -0.12  0.85 | 0.01  0.45 | -0.11  0.86 |
| **Anti-TP**  **(% _positives_)** | **β**  **p-val** | -0.02  0.98 | -0.01  0.93 | 0.01  0.88 | 0.002  0.96 | -0.94  0.28 | 0.74  0.41 | -0.01  0.52 | -1.43  0.11 |
| **Anti-TG**  **(% _positives_)** | **β**  **p-val** | 1.05  0.40 | -0.03  0.64 | -0.02  0.63 | 0.002  0.97 | 18.10  1.00 | -0.43  0.71 | -0.01  0.53 | -1.98  0.13 |
| **ABGAs**  **(% _positives_)** | **β**  **p-val** | -0.63  0.32 | -0.03  0.45 | -0.03  0.26 | -0.05  0.12 | 1.40  0.20 | -1.27  0.24 | 0.01  0.07 | 0.83  0.45 |

**Legend**: β – unstandardized coefficient for the variable immune marker; ABGAs – antibasal ganglia antibodies; AD – antidepressant; AID – autoimmune-disorder; ANAs – anti-nuclear antibodies; Dep. Episode – current depressive episode; IL – interleukin; IFN – interferon; OCD – obsessive-compulsive disorder; STAI – state-trait anxiety inventory; TG – thyroglobulin; TGF – transforming growth factor; TNF – tumor necrosis factor; TP – thyroid peroxidase; YBOCS – Yale Brown Obsessive-Compulsive Scale. ^1^Anholt et al. Psychol Med 2014. ^#^ p-value did not remain significant after controlling for multiple testing using Benjamini-Hochberg’s procedure.
