## Supplementary Table 6 for "Immune (dys)function in obsessive-compulsive disorder"

**Supplementary Table 6** – Primer sequences

| **Marker** | **Sequence (5’->3’)** | **Length** | **Tm** | **GC %** | **Self complementarity** | **Self 3’ complementarity** | **Product length (bp)** | **Reference** |
| --- | --- | --- | --- | --- | --- | --- | --- | --- |
| **IL-1β**  Forward  Reverse | ACAGATGAAGTGCTCCTTCCA  GTCGGAGATTCGTAGCTGGAT | 21  21 | 59.02  59.38 | 47.62  52.38 | 4.00  4.00 | 2.00  2.00 | 73 | Li et al. Human Molecular Genetics, 2004 |
| **IL-2**  Forward  Reverse | AACTCACCAGGATGCTCACATTTA  TCCCTGGGTCTTAAGTGAAAGTTT | 24  24 | 60.26  59.83 | 41.67  41.67 | 4.00  6.00 | 2.00  5.00 | 148 | Overbergh et al. Journal of Biomolecular Techniques, 2003 |
| **IL-4**  Forward  Reverse | CCACGGACACAAGTGCGATA  CCCTGCAGAAGGTTTCCTTCT | 20  21 | 60.39  59.93 | 55.00  52.38 | 4.00  9.00 | 2.00  9.00 | 101 | Overbergh et al. Journal of Biomolecular Techniques, 2003 |
| **IL-6**  Forward  Reverse | TGAACTCCTTCTCCACAAGCG  TCTGAAGAGGTGAGTGGCTGTC | 21  21 | 60.27  61.66 | 52.38  54.55 | 4.00  4.00 | 2.00  1.00 | 151 | Tchirkov et al. British Journal of Cancer, 2001 |
| **IL-8**  Forward  Reverse | CTTGGCAGCCTTCCTGATTT  TTCTTTAGCACTCCTTGGCAAAA | 20  23 | 58.45  59.04 | 50.00  39.13 | 5.00  5.00 | 0.00  2.00 | 67 | Kamsteeg et al. The American Journal of Pathology, 2011 |
| **IL-10**  Forward  Reverse | GTGATGCCCCAAGCTGAGA  CACGGCCTTGCTCTTGTTTT | 19  20 | 59.70  59.61 | 57.89  50.00 | 4.00  6.00 | 1.00  0.00 | 138 | Overbergh et al. Journal of Biomolecular Techniques, 2003 |
| **IL-12p40**  Forward  Reverse | TGGAGTGCCAGGAGGACAGT  TCTTGGGTGGGTCAGGTTTG | 20  20 | 62.68  59.82 | 60.00  55.00 | 3.00  2.00 | 3.00  0.00 | 147 | Overbergh et al. Journal of Biomolecular Techniques, 2003 |
| **IL-17α**  Forward  Reverse | TCCCACGAAATCCAGGATGC  GGATGTTCAGGTTGACCATCAC | 20  22 | 60.11  59.25 | 55.00  50.00 | 7.00  9.00 | 3.00  7.00 | 75 | Li et al. Human Molecular Genetics, 2004 |
| **IL-23**  Forward  Reverse | TCTCCTTCTCCGCTTCAAAATC  GGCGGCTACAGCCACAAA | 22  18 | 58.40  60.67 | 45.45  61.11 | 3.00  6.00 | 1.00  0.00 | 58 | Li et al. Human Molecular Genetics 2004 |
| **IFN-γ**  Forward  Reverse | TCAGCTCTGCATCGTTTTGG  TCAGCTCTGCATCGTTTTGG | 20  24 | 58.84  58.70 | 50.00  41.67 | 4.00  3.00 | 0.00  3.00 | 120 | Overbergh et al. Journal of Biomolecular Techniques, 2003 |
| **TGF-β**  Forward  Reverse | CAGCAACAATTCCTGGCGATA  AAGGCGAAAGCCCTCAATTT | 21  20 | 58.98  58.37 | 47.62  45.00 | 5.00  4.00 | 2.00  3.00 | 136 | Overbergh et al. Journal of Biomolecular Techniques, 2003 |
| **TNF-α**  Forward  Reverse | TGCTCCTCACCCACACCAT  GGAGGTTGACCTTGGTCTGGTA | 19  22 | 60.85  61.36 | 57.89  54.55 | 2.00  6.00 | 2.00  2.00 | 60 | Li et al. Cell Metabolism, 2015 |
| **GAPDH**  Forward  Reverse | TCCAAAATCAAGTGGGGCGA  TGATGACCCTTTTGGCTCCC | 20  20 | 59.89  59.96 | 50.00  55.00 | 3.00  3.00 | 0.00  1.00 | 115 | Xiao et al. Peer J, 2017 |
| **β2-MG**  Forward  Reverse | TGGGTTTCATCCATCCGACA  TCAGTGGGGGTGAATTCAGTG | 20  21 | 59.01  59.93 | 50.00  52.38 | 3.00  8.00 | 0.00  3.00 | 138 | Xiao et al. Peer J, 2017 |
| **Β-actin**  Forward  Reverse | GGCACCCAGCACAATGAAG  CCGATCCACACGGAGTACTTG | 19  21 | 59.41  60.47 | 57.89  57.14 | 3.00  6.00 | 0.00  3.00 | 66 | Valente et al. BMC Molecular Biology, 2009 |

**Legend**: β2-MG – β2-microglogulin; GAPDH - glyceraldehyde 3-phosphate dehydrogenase; IL – interleukin; TGF – transforming growth factor; TNF – tumour necrosis factor.
